# Provider Density and Systemic Contributors to Rural Cardiovascular Disease Mortality in New England

**DOI:** 10.1101/2025.01.29.25321338

**Authors:** Eashwar Krishna

**Affiliations:** Department of Sociology, University of Connecticut, Storrs, Connecticut

**Keywords:** Rural hospitals, rural communities, New England, cardiologists, heart failure

## Abstract

**Purpose:** Rural communities face substantial challenges in accessing healthcare, which contribute to worse outcomes, particularly among patients with chronic heart disease. Rural disparities in cardiovascular disease (CVD)-related mortality may be compounded by limited access to primary care and specialist providers. This study aims to quantify the relationship between CVD-related mortality and the availability of primary-care (PCP) and specialist providers in rural and non-rural areas of New England.

**Methods:** This study examined associations between county-level provider density and mortality rates for inpatient heart failure (HF) at 154 New England hospitals and heart attacks (HA) at 99 New England hospitals. I assessed rural/non-rural differences in provider densities and mortality using t-tests. I used multivariable linear models to regress mortality rates onto rurality, provider density, and socioeconomic variables.

**Results:** Rural spaces had a lower density of cardiologists (*B*=–0.522, 95% *CI* [–0.809, –0.234], *p*<.001) but not PCP density (*B*=–0.008, 95% *CI* [–0.313, 0.304], *p*=.973). Rurality was also associated with higher 30-day inpatient HF mortality rates after controlling for covariates (*B*=0.492, 95% *CI* [0.185, 0.828], *p*=.005); this was not true for inpatient HA mortality (*B*=0.348, 95% *CI* [–0.416, 1.005], *p*=.266). Cardiologist density was significantly associated with HF mortality in unadjusted models (*B*=–0.398, 95% *CI* [–0.586, –0.208], *p*<.001) but not adjusted (*B*=–0.079, 95% *CI* [–0.348, 0.162], *p*=.479).

**Conclusion:** There exists an intersection between socioeconomic gradients and rurality, underscoring the importance of addressing structural inequities to reduce rural health disparities. Targeted innovations in the recruitment and support of specialist physicians in rural spaces would reduce heart failure mortality.

## 1. INTRODUCTION

Rural communities face substantial challenges in accessing consistent healthcare, contributing to worse outcomes, particularly among patients with chronic diseases. The prevalence of chronic heart disease (CHD) is notably higher in rural populations, affecting 14.2% of residents compared to 11.2% in small metropolitan areas and 9.9% in urban settings.^1^ Moreover, rural patients presenting with acute myocardial infarction and heart failure in various regions of the world have higher mortality rates, and longitudinal analyses have demonstrated a significantly higher cardiovascular disease mortality rate in rural counties than urban counties.^2,3^

These disparities are compounded by limited access to timely follow-up care and low provider density, which are crucial for managing conditions like congestive heart failure (CHF). For example, early follow-up within seven days of discharge has been shown to reduce 30-day readmission rates for CHF patients, a critical factor in improving outcomes.^4^ Unfortunately, rural areas often lack a sufficient number of specialists and primary care providers (PCPs) alike, making such follow-up difficult. It is also known that specialist follow up tends to be associated with lower mortality after CHF diagnosis when compared to general medicine follow-ups or none at all.^5,6^

While most individuals regardless of rural status live within twenty kilometers (i.e., about 12 miles) of a primary care provider, accessing heart failure cardiologists can require rural patients to travel over one hundred kilometers (i.e., about 62 miles), a burden few can manage without substantial resources^7^; rural adults with congenital heart diseases frequently identified distance from the clinic as a barrier to seeking care, much more so than their urban counterparts.^8^ As such, higher provider density in a particular geographic area is correlated with higher rates of early follow-up.^4^ For many rural physicians themselves, the challenges don’t end with geography. They often face overwhelming patient loads, an outcome caused by the low number of physicians sharing the burden, leading to burnout and high turnover rates. Moteirak et al. showed that many new providers find it difficult to sustain their practice amid such demands and commonly leave after one or two years.^7^ This constant cycle of provider shortages and turnover exacerbates the challenges of delivering consistent, high-quality care in rural areas, particularly for chronic conditions requiring specialized follow-up.

Disparities in inpatient mortality between rural and urban hospitals are less definitive and often differ between studies. Minhas et. al. examined trends in the National Inpatient Sample from 2004 to 2018 and discovered that urban hospitals had a lower inpatient mortality rate for heart failure (HF) and acute myocardial infarction (AMI) patients; however, the authors also noted that the mortality gap consistently decreased over the study period, indicating an improvement in rural specialist care over time.^9^ A similar paper by Patlolla et. al. seems to report almost contradictory results: an analysis of the same database from 2000 to 2017 revealed higher inpatient mortality rates for AMI patients with cardiac arrest complications in urban hospitals when compared to rural hospitals.^10^

Despite extensive research documenting rural-urban disparities in healthcare outcomes, there remains a lack of county-level analysis on how provider density is associated with inpatient cardiovascular disease (CVD) outcomes. Research has demonstrated that outpatient resources like disease management clinics (DMCs) staffed by outpatient cardiologists significantly lower all-cause mortality following hospitalizations for HF, providing substantial basis to the hypothesized connection in this study.^11^ The leading analysis of provider density and associated mortality rates comes from a 2013 analysis of claims data to examine post-hospitalization 30-day mortality risk for AMI and HF patients.^12^ While the authors did not explicitly examine rurality, they conducted regional analysis using hospital referral regions (HRRs), which are larger units that reflect care patterns for specialist interactions^13^; these typically span multiple counties.

These HRRs were organized into quintiles based on cardiologist density, and patients from the lowest quintile HRRs had a modestly higher 30-day mortality risk as compared to patients in the highest quintile. This study provided strong evidence for the impact of specialist density on patient outcomes; however, the usage of HRRs did not allow rural-urban comparison, as the authors note that these regions often include both kinds of areas. Particularly in a region such as New England, which contains many small-area rural communities, an HRR-level analysis may not capture the true impact of rurality. Rather, a county-level analysis may provide a more nuanced perspective into the issue. Furthermore, the usage of quintiles may not reveal as clear of an association as a linear regression might.

Studies such as one conducted by Harris et. al indicate that in particular regions, socioeconomic factors such as poverty, rather than rurality or provider density considered in isolation, primarily drive hospitalization rates.^14^ In addition, income level and educational attainment are significantly associated with CVD rates in high-income countries such as the U.S, demanding region-specific analysis to elucidate potential varying contributions of socioeconomic variables.^15^ As such, there appears to be a multifaceted influence of rurality and other social determinants of health on cardiovascular disease prevalence, subsequent hospitalization, and thus inpatient mortality rates.

## 2. METHODS

### 2.1 Data Sources

Thirty-day inpatient mortality rates coded as heart failure or myocardial infarction were obtained from the National Complications and Deaths dataset published by the Centers for Medicare and Medicaid Services (CMS) on January 31st, 2024, which reported these measures from July 1st, 2019, to June 30th, 2022 for hospitals in all U.S states.^16^ The usage of a multi-year dataset increases the sample size with regard to the patient pool and therefore makes the measure more resilient against chance fluctuations; similar studies such as Krumholz et. al. have used a 3-year window.^17^ This dataset was filtered to exclude all data points except those located in Connecticut, Rhode Island, Massachusetts, New Hampshire, Vermont, and Maine.^18^ Of the 182 hospitals included in the study, 28 did not have data for heart failure mortality rates, and 83 did not have data for heart attack mortality; in both cases, this indicated that fewer than twenty-five deaths from either condition had occurred in the three-year timeframe. Patients were excluded from CMS calculations of heart failure (HF) and heart attack (HA) mortality rates if they were: (a) diagnosed with COVID-19 upon admission, (b) had inconsistent/unknown demographics or vitals, (c) discharged against medical advice, (d) discharged alive on the same day or day-after hospital admission, or (e) enrolled in Medicare hospice at any point in the year prior to admission. Furthermore, patients with a left ventricular assist device (LVAD) implementation or those who received a heart transplant up to twelve months before admission were excluded from heart failure calculations.

Hospitals were coded as rural or non-rural using the Rural Health Grants Eligibility tool based on the Federal Office of Rural Health Policy definition, which uses a modified interpretation of Rural-Urban Commuting Area codes to ensure representation of rural tracts that are within a nominally urban or suburban county.^19^ The number of primary care and cardiology care providers by county was drawn from the Health Resources and Services Administration (HRSA) Area Health Resource files 2022–2023 release, which included data up to 2021.^20^

Provider density (i.e., practitioners per ten thousand residents) was calculated using county population data from the 2022 American Community Survey (ACS) release.^21^

Percentage of residents with a college degree, and percentage of residents below the poverty line were measured in 2021; median age was measured in 2022. Values were drawn from the County Health Rankings as published by the University of Wisconsin Population Health Institute,^22^ Federal Reserve Economic Data as released by the St. Louis Federal Reserve,^23^ the 2022 ACS release,^21^ and the Small Area Income and Poverty Estimates released by the U.S Census Bureau.^24^

### 2.2 Statistical Analysis

A combination of descriptive, bivariate, and multivariable regression analyses was performed to evaluate the relationship between structural factors, rurality, provider density, and mortality outcomes. Two-tailed t-tests were utilized to compare the key predictors and outcomes; equal variances were not assumed. Linear regressions were employed to test the strength of the interactions; all variables except rurality (i.e., a binary value where rural=1) were Z-normalized. Standardized coefficients (*B*) represent the change in outcome in standard deviation units (*SDU*) per increase in one *SD* of the predictor. To ensure robust interval estimation, bias-corrected and accelerated (*BCa*) 95% confidence intervals (*CIs*) were calculated using 5,000 bootstrap resamples. For heart attack (HA) mortality and heart failure (HF) mortality, two sets of regression analyses were conducted to compare rural and non-rural hospitals: Models 1 and 3, respectively, did not control for covariates, while Models 2 and 4 adjust for county-level percentage of residents with a college degree, median age, and percentage of residents below the poverty line. The decision to retain multiple models in the final reporting for mortality outcomes stemmed from the understanding that rural disparities do not exist in a vacuum but rather in synergy with social determinants of health. As such Models 1 and 3 demonstrate the disparity “as-is” whereas Models 2 and 4 display the isolated association with rurality, which is by nature a more theoretical measure. All analyses were conducted using IBM SPSS Statistics Version 30 with a significance threshold of α=0.05.

## 3. RESULTS

Table 1 reports descriptive statistics for study variables from New England hospitals and their county locations. Rural hospitals in New England demonstrated significantly higher 30-day inpatient mortality rates for heart failure (12.37 vs. 10.80; *p*<.001) but no significant difference in heart attack mortality compared to non-rural hospitals during 2019–2022 (12.80 vs. 12.22; *p*=.122). Rural counties exhibited significantly fewer cardiologists per 10,000 residents (0.74 vs. 1.35, *p*<.001), though primary care provider density did not differ significantly between rural and non-rural areas (9.85 vs. 9.88; *p*=.960). Rural populations were older (median age of 46.31 vs. 40.00; *p*<.001) and less likely to have a college degree (33.21% vs. 41.82%; *p*<.001). Poverty rates showed no significant variation (11.00% vs. 11.19%; *p*=.692).

**Table 1.** Descriptive Statistics for Study Variables from New England Counties and <u>Hospitals.</u>.

| *Measures* | Rural |  | Non-Rural |  |
| --- | --- | --- | --- | --- |
|  | *Mean* | *SD* | *Mean* | *SD* |
| *Outcome Measures* |  |  |  |  |
| Inpatient 30-day Heart Failure Mortality Rate<sup>a</sup> | 12.37*** | (1.65) | 10.80 | (1.72) |
| Inpatient 30-day Heart Attack Mortality Rate<sup>b</sup> | 12.80* | (1.31) | 12.22 | (1.02) |
| *Key Measures of Interest<sup>c</sup>* |  |  |  |  |
| Primary Care Providers per 10,000 residents | 9.85 | (3.85) | 9.88 | (3.12) |
| Cardiologists per 10,000 residents | 0.74*** | (1.11) | 1.35 | (1.16) |
| *Controls* |  |  |  |  |
| Percent below the Poverty Line | 11.00 | (2.76) | 11.19 | (3.82) |
| Percent with a College Degree | 33.21*** | (8.71) | 41.82 | (9.29) |
| Median Age | 46.31*** | (3.22) | 40.00 | (3.57) |

<sup>a</sup> 52 rural hospitals and 102 non-rural hospitals reported this measure from 2019-2022.
<sup>b</sup> 15 rural hospitals and 84 non-rural hospitals reported this measure from 2019-2022
<sup>c</sup> 35 rural counties and 22 non-rural counties reported these measures from 2021-2022
\* $p < .05$ ; \*\* $p < .01$ ; \*\*\* $p < .001$ (two-tailed tests).

Table 2 shows regressions estimating heart attack mortality, and heart failure mortality for New England counties and hospitals. Model 1 showed no significant association between rurality and primary care provider density (*B*=−0.008; *95% CI*: [−0.313, 0.304]; *p*=.973), but Model 2 revealed that rural have lower cardiologist density (*B*=−0.522 [−0.809, −0.234]; *p*<.001). In Models 1 and 2, rurality was not associated heart attack mortality in unadjusted models (*B*=0.537 [−0.117, 1.167]; *p*=.055) or adjusted models (*B*=0.348 [−0.416, 1.005]; *p*=.266). Model 1 identifies that a standard deviation increase in cardiology density is associated with a.277 reduction in heart attack mortality. In Model 2, the direction and strength remains similar, but the association does not reach significance.

**Table 2.** Linear Regressions Estimating Heart Attack Mortality, and Heart Failure Mortality for New England Counties and Hospitals.

|  | Inpatient 30-day<br>Heart Attack<br>Mortality Rate |  | Inpatient 30-day<br>Heart Failure<br>Mortality Rate |  |
| --- | --- | --- | --- | --- |
| *Predictors* | *Model 1* | *Model 2* | *Model 3* | *Model 4* |
| Rural (yes=1) | 0.537<br>[−0.117,<br>1.167] | 0.348<br>[−0.416,<br>1.005] | 0.895***<br>[0.619,<br>1.178] | 0.492**<br>[0.185,<br>0.828] |
| *Provider Density* |  |  |  |  |
| Primary Care<br>Provider<br>Density | −0.131<br>[−0.363,<br>0.104] | 0.112<br>[−0.154, 0.419] | −0.167*<br>[−0.358,<br>0.032] | 0.134<br>[−0.065,<br>0.362] |
| Cardiology<br>Density | −0.277*<br>[−0.541,<br>−0.021] | −0.262<br>[−0.678,<br>0.193] | −0.398***<br>[−0.586,<br>−0.208] | −0.079<br>[−0.348,<br>0.162] |
| Sample size | 99 | 99 | 154 | 154 |

*Note:* Standardized coefficients presented (*B*) with 95% confidence intervals in brackets. Models 1 and 3 do not control for covariates, and Models 2 and 4 control for but do not show median age, percent below the poverty line, and percent with a college degree. Sample sizes differ across models due to censored information in cases where fewer than 25 patients in a hospital experienced mortality due to heart attack or heart failure. Mortality rates are measured over the period 2019-2022; provider density and control variables are from 2020-2021.
\* $p < .05$ ; \*\* $p < .01$ ; \*\*\* $p < .001$ (two-tailed tests).

Model 3 shows that rurality was associated with significantly higher heart failure mortality risk in the unadjusted equation (*B*=0.895; [0.619, 1.178]; *p*<.001). Likewise, counties with greater density of primary care providers (*B*=−0.167; [−0.358, 0.032]; *p*=.044) and cardiologists (*B*=−0.398; [−0.586, −0.208]; *p*<.001) have lower heart attack mortality rates, on average. In Model 4, the equation adjusts for socioeconomic measures and identifies a similarly higher rate of heart failure mortality in rural counties (*B*=0.492; [0.185, 0.828]; *p*=.005). The significant associations in unadjusted models between primary care density (*B*=0.134; [−0.065, 0.362]; *p*=.140), cardiologist density (*B*=−0.079; [−0.348, 0.162]; *p*=.479), and heart failure mortality are no longer statistically significant in Model 4.

## 4. DISCUSSION

This study observed significant differences in healthcare access and outcomes between rural and non-rural areas, particularly in heart failure mortality. Rural hospitals have substantially higher heart failure mortality rates compared to urban hospitals, but this disparity lessens when accounting for socioeconomic factors such as poverty and educational attainment. This finding suggests that disparities in social determinants of health (SDOH) play a central role in shaping rural-urban inequities, echoing evidence from other contexts that suggests rural areas perform poorly in multiple SDOH metrics.^25^ Notably, no significant difference was observed in heart attack mortality rates between rural and urban hospitals, which may reflect the acute, standardized nature of heart attack treatment protocols that are less dependent on long-term access to specialist care.^26^ This contrast between conditions requiring ongoing management, such as heart failure, and acute events, like heart attacks, is a lesson in the importance of examining disease-specific drivers of rural-urban disparities.

While covariates partially account for the rural-urban disparity in cardiologist density, they do not fully explain it. The structural challenges faced by rural hospitals—geographic isolation, limited financial resources, and reduced access to professional development opportunities—are likely significant contributors to these disparities.^27^ It may also be the case that there is a financial barrier to placing specialists in community settings parallel to general practitioners.^28^ Cardiologist density was significantly associated with heart failure mortality in unadjusted analyses, and remained so even after adjusting for confounders albeit the size of the effect was somewhat reduced. This suggests that while specialist availability is critical, its impact and perhaps the variable itself are mediated by structural factors. For heart attack mortality, the absence of significant associations with cardiologist density may reflect the effectiveness of the aforementioned standardized treatment protocols, which reduce variability in care delivery across settings and allow non-physician providers and general practitioners to fill the gaps.

In contrast to cardiologist density, PCP density showed no significant difference between rural/non-rural areas. Educational attainment emerged as a critical determinant of provider distribution: counties with higher college-educated populations had significantly more PCPs and cardiologists. Furthermore, the lack of a strong association between PCP density and heart failure or heart attack mortality reinforces the idea that primary care providers, while essential for general health maintenance, may not be as directly impactful on outcomes for conditions requiring specialist intervention. This highlights a potential gap in the rural healthcare infrastructure: even when primary care access is comparable, the lack of specialists may leave rural populations underserved for complex or chronic conditions like heart failure.

The role of socioeconomic variables in shaping healthcare outcomes is evident across the analyses. Education and poverty consistently influenced provider availability and mortality rates. Lower-income populations, for instance, have been reported to have increased cardiovascular disease hospitalization due to cost-related medication underuse,^29^ which likely contributes to outcome disparities in rural areas. Higher education was consistently associated with improved outcomes and provider density, emphasizing the need for policies targeting educational and economic inequities in rural regions.

Although these socioeconomic trends account for a non-negligible portion of the variance in provider density and health outcomes, they do not fully explain the structural inequities that persist in rural healthcare systems. Conclusions from previous literature are mixed, with some noting that quality and access to care remains a significant factor beyond SDOH metrics in rural outcome disparities^30^; while others report that controlling for social risk factors completely attenuated rural-urban disparities in CHD.^31^ Taken together, these results highlight the multifaceted nature of the issue and suggest that regional-level, rather than national-level, analysis may be more informative.

### 4.1 Implications

The need for a targeted approach raises several critical considerations for rural health policy, particularly in addressing the disparities in cardiologist access. One promising avenue is the implementation of recruitment and retention programs tailored specifically for rural areas. Programs such as the Physicians for Shortage Area Program (PSAP) at Jefferson Medical College provide a useful model for this approach. PSAP has demonstrated substantial success in increasing the percentage of rural family physicians in Pennsylvania, accounting for a significant portion of the rural physician workforce despite the program’s relatively small size.^32^ More broadly speaking, any such efforts must address both geographic and socioeconomic barriers. For example, incentivizing specialists to practice in low-income rural counties through loan forgiveness tied to local income levels could mitigate dual inequities. Similarly, expanding telehealth to bridge geographic gaps should be paired with programs to improve broadband access and digital literacy in underserved communities

The introduction of similar programs targeting cardiologists and other specialists, coupled with incentives such as rural residency rotations and preferences for in-state applicants, could play a pivotal role in building a more sustainable rural healthcare workforce. It follows from logic that the providers most likely to practice in rural areas are those who have lived or trained in such,^33^ and medical education institutions seeking to produce rural physicians will likely see success by creating more rural training opportunities, as positive exposure to rural environments during medical training is a known retention factor.^34^ Numerous initiatives currently aim to attract physicians to underserved regions with a disproportionate emphasis on recruitment, with less attention given to retaining these providers after their initial service commitments are met.^35^ Furthermore, the vast majority of newly-created rural residency programs have a family/internal medicine focus^36^; this may reflect a crippling oversight to train rural specialists. It should also be noted that such incentive programs were the least prevalent in the northeastern U.S, which includes the New England region studied by the present analysis.^35^

Most existing programs also fail to address the underlying challenges that lead to high turnover rates among rural physicians. Addressing factors such as professional isolation, limited career advancement opportunities, and inadequate support staff, could help build a more stable rural healthcare workforce.^37^ It is imperative that new policy initiatives focus on retention as well as retainment of specialist providers in rural areas; such initiatives would not only help ameliorate the observed health disparities, but would also increase the social integration of specialists within rural areas, which may have the additional beneficial effect of increasing trust in the medical establishment among these communities.

Another key policy implication is the need for telemedicine and outreach programs as short-term solutions to bridge the gap in access to specialist care. Rural hospitals and clinics often face significant challenges in recruiting cardiologists due to geographic isolation and limited financial resources, but telemedicine provides an opportunity to extend the reach of urban-based specialists to underserved areas. By enabling remote consultations and follow-up care, telemedicine can reduce the need for rural patients to travel long distances for specialist appointments, which is a significant barrier to care in these settings.^38^ This approach is particularly well-suited to managing chronic conditions like heart failure, where regular monitoring and adjustments to treatment plans are critical. Telemedicine has already shown promise in improving cardiovascular care in rural communities and appears to be well-received by rural patients.^39–41^

However, telemedicine alone is not sufficient to address the long-term needs of rural populations. While it offers a mechanism to improve access, it cannot fully substitute for the presence of specialists within these communities. Policies should therefore support innovative care models such as visiting consultant clinics,^42^ where urban-based cardiologists provide on-site care in rural hospitals and clinics on a rotating basis. These clinics not only bring specialist expertise directly to rural areas but also facilitate collaboration between primary care providers and specialists, which can enhance the overall quality of care. The combination of telemedicine and visiting consultant clinics offers a more comprehensive strategy for addressing the immediate and ongoing needs of rural patients.

This study contributes to this body of research by quantifying the relationships between provider density, mortality outcomes, and socioeconomic factors in the New England region, a geographic area that has received less specific attention in rural health research compared to other parts of the United States. Among the existing literature, this paper seeks to extend knowledge on how county-level specialist density is linked to mortality outcomes in the rural context.

### 4.2 Limitations

One notable limitation in this analysis is the potential for missing data among hospitals with low mortality counts, particularly for heart attack patients, which could lead to an underrepresentation of smaller facilities in the analysis. Such smaller facilities are more likely to be in rural tracts^43^; the implication for this study is that while HF mortality disparities are likely satisfactorily captured, the same for HA may not have been, leading to insignificant results in that metric. Post-hoc analysis options of statistical power are themselves limited by the lack of data available to establish baseline expectations. Additionally, unmeasured confounders, such as differences in hospital resources, staffing levels, or patient comorbidities, may have influenced the results. While the inclusion of income, education, and age provides a robust framework for isolating the impact of rurality, it is possible that other variables not accounted for in the models, such as hospital metrics and local telehealth regulations, could also play a role in explaining the observed disparities. Future research should aim to address these gaps by incorporating more comprehensive data and exploring the impact of other potential confounders on rural-urban health inequities.

The finding that primary care providers are relatively equitably distributed across New England counties should not be taken out of the context of the metric used; the fact that there are equal rates of providers per ten thousand residents in rural and non-rural counties is not sufficient to demonstrate equitable access. It is also true that certain rural counties, most notably those in the northern regions of Maine, are much larger in terms of land area while simultaneously being sparsely populated. As such, the rate of providers by population may seem sufficient, yet the actual access to regular health care is still inhibited by other, uniquely rural, factors such as geography. Indeed, distance to primary care providers has been isolated as a significant contributor to higher disease burdens.^44^

Furthermore, the results of these analyses should not be interpreted as diminishing the irrefutable impact of primary care providers on preventing and managing chronic cardiovascular conditions. This study was limited by its usage of inpatient mortality, which reflects the relative progression of patients into severe statuses and the management of said status at complex stages.

As such, there exists the chance that the analysis overemphasized the role of specialist care, considering that inpatient care is the setting in which specialist care is the most critical. Alternatively, outpatient mortality could be utilized, yet further structural factors such as EMS response systems would need to be controlled for.

Despite these limitations, the findings offer valuable insights into the systemic and structural challenges facing rural healthcare systems. They highlight the urgent need for targeted interventions to improve access to specialist care, particularly for conditions like heart failure that require ongoing management and coordination. Policymakers should consider the implementation of comprehensive strategies that address both the immediate and long-term needs of rural communities, including telemedicine expansion, visiting consultant clinics, and programs to recruit and retain specialists. By addressing the multifaceted nature of rural healthcare inequities, these policies have the potential to significantly improve health outcomes for underserved populations.

## 5. CONCLUSION

Rural hospitals in New England exhibit higher heart failure mortality rates compared to non-rural hospitals, a disparity that diminishes but does not fully disappear when adjusting for socioeconomic factors such as education and poverty. The shortage of cardiologists in rural areas stands out as a key contributor, emphasizing the structural challenges these communities face in accessing specialized care. Primary care provider density, which shows no significant urban-rural difference, further suggests that rural cardiovascular-health-inequities are driven more by specialized care shortages than general healthcare access. This analysis underscores the urgent need for targeted interventions to improve rural health outcomes and promote healthcare equity.

## Data Availability

All data produced in the present study are available upon reasonable request to the authors

## Acknowledgements

The author would like to thank Dr. Ryan Talbert Department of Sociology, University of Connecticut for his guidance in the writing of this manuscript

## Notes

Disclosures: The author has no conflicts of interest to disclose

### Competing Interest Statement

The authors have declared no competing interest.

### Funding Statement

This study did not receive any funding

### Summary of Updates

Revisions were made to enhance the stability of regression models by re-orienting covariates.

## References

1. Schopfer DW. Rural health disparities in chronic heart disease. Prev Med. 2021;152:106782. doi:10.1016/j.ypmed.2021.106782

2. Faridi B, Davies S, Narendrula R, et al. Rural–urban disparities in mortality of patients with acute myocardial infarction and heart failure: a systematic review and meta-analysis. Eur J Prev Cardiol. 2025;32(4):327–335. doi:10.1093/eurjpc/zwae351

3. Son H, Zhang D, Shen Y, et al. Social Determinants of Cardiovascular Health: A Longitudinal Analysis of Cardiovascular Disease Mortality in US Counties From 2009 to 2018. J Am Heart Assoc. 2023;12(2):e026940. doi:10.1161/JAHA.122.026940

4. Verdejo HE, Ferreccio C, Castro PF. Heart Failure in Rural Communities. Heart Fail Clin. 2015;11(4):515–522. doi:10.1016/j.hfc.2015.07.011

5. Indridason OS, Coffman CJ, Oddone EZ. Is specialty care associated with improved survival of patients with congestive heart failure? Am Heart J. 2003;145(2):300–309. doi:10.1067/mhj.2003.54

6. Abubakar I, Kanka D, Arch B, Porter J, Weissberg P. Outcome after acute myocardial infarction: a comparison of patients seen by cardiologists and general physicians. BMC Cardiovasc Disord. 2004;4:14. doi:10.1186/1471-2261-4-14

7. Motairek I, Chen Z, Makhlouf MHE, et al. Mapping Geographic Proximity to Cardiologists Across the United States. Circ Cardiovasc Qual Outcomes. 2023;16(10). doi:10.1161/CIRCOUTCOMES.123.010133

8. McGrath L, Taunton M, Levy S, Kovacs AH, Broberg C, Khan A. Barriers to care in urban and rural dwelling adults with congenital heart disease. Cardiol Young. 2022;32(4):612–617. doi:10.1017/S1047951121002766

9. Minhas AMK, Sheikh AB, Ijaz SH, et al. Rural-Urban Disparities in Heart Failure and Acute Myocardial Infarction Hospitalizations. Am J Cardiol. 2022;175:164–169. doi:10.1016/j.amjcard.2022.04.014

10. Patlolla SH, Pajjuru VS, Sundaragiri PR, et al. Hospital-Level Disparities in the Management and Outcomes of Cardiac Arrest Complicating Acute Myocardial Infarction. Am J Cardiol. 2022;169:24–31. doi:10.1016/j.amjcard.2021.12.057

11. Van Spall HGC, Rahman T, Mytton O, et al. Comparative effectiveness of transitional care services in patients discharged from the hospital with heart failure: a systematic review and network meta‐analysis. Eur J Heart Fail. 2017;19(11):1427–1443. doi:10.1002/ejhf.765

12. Kulkarni VT, Ross JS, Wang Y, et al. Regional density of cardiologists and rates of mortality for acute myocardial infarction and heart failure. Circ Cardiovasc Qual Outcomes. 2013;6(3):352–359. doi:10.1161/CIRCOUTCOMES.113.000214

13. Appendix on the Geography of Health Care in the United States. Published online 1996. https://data.dartmouthatlas.org/downloads/methods/geogappdx.pdf

14. Harris DE, Aboueissa AM, Hartley D. Myocardial infarction and heart failure hospitalization rates in Maine, USA - variability along the urban-rural continuum. Rural Remote Health. 2008;8(2):980.

15. Schultz WM, Kelli HM, Lisko JC, et al. Socioeconomic Status and Cardiovascular Outcomes: Challenges and Interventions. Circulation. 2018;137(20):2166–2178. doi:10.1161/CIRCULATIONAHA.117.029652

16. Centers for Medicare and Medicaid Services. Complications and Deaths - National. Published online 2024. https://data.cms.gov/provider-data/dataset/qqw3-t4ie#data-table

17. Krumholz HM, Lin Z, Keenan PS, et al. Relationship Between Hospital Readmission and Mortality Rates for Patients Hospitalized With Acute Myocardial Infarction, Heart Failure, or Pneumonia. JAMA. 2013;309(6):587. doi:10.1001/jama.2013.333

18. Census Regions and Divisions of the United States. Published online May 21, 2025. https://www2.census.gov/geo/pdfs/maps-data/maps/reference/us_regdiv.pdf

19. Health Resources and Services Administration. Rural Health Grants Eligibility Analyzer. Published online 2020. https://data.hrsa.gov/tools/rural-health

20. Health Resources and Services Administration. Area Health Resources Files. Published online July 31, 2023. https://data.hrsa.gov/topics/health-workforce/ahrf

21. U.S Census Bureau. American Community Survey 5-year estimates. Published online 2022. https://data.census.gov/table/ACSST1Y2022.S0101

22. University of Wisconsin Population Helath Institute. County Health Rankings & Roadmaps. Published online 2022. https://www.countyhealthrankings.org/health-data/health-factors/social-economic-factors/income/

23. St. Louis Federal Reserve. Federal Reserve Economic Data. Published online 2021. https://fred.stlouisfed.org/release/tables?rid=330&eid=391810

24. U.S Census Bureau. Small Area Income and Poverty Estimates (SAIPE) Program. Published online October 5, 2022. https://www.census.gov/programs-surveys/saipe.html

25. Weeks WB, Chang JE, Pagán JA, et al. Rural-urban disparities in health outcomes, clinical care, health behaviors, and social determinants of health and an action-oriented, dynamic tool for visualizing them. Wang Z, ed. PLOS Glob Public Health. 2023;3(10):e0002420. doi:10.1371/journal.pgph.0002420

26. Franklin C, Mathew J. Developing strategies to prevent inhospital cardiac arrest: analyzing responses of physicians and nurses in the hours before the event. Crit Care Med. 1994;22(2):244–247.

27. Collins C. Challenges of Recruitment and Retention in Rural Areas. N C Med J. 2016;77(2):99–101. doi:10.18043/ncm.77.2.99

28. Lovén M, Pitkänen LJ, Paananen M, Torkki P. Evidence on bringing specialised care to the primary level—effects on the Quadruple Aim and cost-effectiveness: a systematic review. BMC Health Serv Res. 2024;24(1):2. doi:10.1186/s12913-023-10159-6

29. Heisler M, Choi H, Rosen AB, et al. Hospitalizations and Deaths Among Adults With Cardiovascular Disease Who Underuse Medications Because of Cost: A Longitudinal Analysis. Med Care. 2010;48(2):87–94. doi:10.1097/MLR.0b013e3181c12e53

30. Zeitler EP, Joly J, Leggett CG, et al. The role of comorbidities, medications, and social determinants of health in understanding urban‐rural outcome differences among patients with heart failure. J Rural Health. 2024;40(2):386–393. doi:10.1111/jrh.12803

31. Liu M, Marinacci LX, Joynt Maddox KE, Wadhera RK. Cardiovascular Health Among Rural and Urban U. Adults—Healthcare, Lifestyle, and Social Factors. JAMA Cardiol. Published online March 31, 2025. doi:10.1001/jamacardio.2025.0538

32. Rabinowitz HK, Diamond JJ, Markham FW, Santana AJ. Increasing the Supply of Rural Family Physicians: Recent Outcomes From Jefferson Medical College’s Physician Shortage Area Program (PSAP): Acad Med. 2011;86(2):264–269. doi:10.1097/ACM.0b013e31820469d6

33. Palnati SR, Jurado CM, Bhakta SH. Exploring Rural Medicine Residency Programs: A Narrative Review. Cureus. Published online October 23, 2024. doi:10.7759/cureus.72238

34. Kumar S, Clancy B. Retention of physicians and surgeons in rural areas—what works? J Public Health. 2021;43(4):e689–e700. doi:10.1093/pubmed/fdaa031

35. Arredondo K, Touchett HN, Khan S, Vincenti M, Watts BV. Current Programs and Incentives to Overcome Rural Physician Shortages in the United States: A Narrative Review. J Gen Intern Med. 2023;38(S3):916–922. doi:10.1007/s11606-023-08122-6

36. Payerchin R. CMS creating residencies to focus on primary care, but they have not expanded to rural areas. Medical Economics. August 14, 2025. https://www.medicaleconomics.com/view/cms-creating-residencies-to-focus-on-primary-care-but-they-have-not-expanded-to-rural-areas

37. Rajagopalan N, Leung SW, Craft RS, Bailey AL. Improving Cardiovascular Health in Rural United States: Role of Academic Medical Centers. JACC Adv. 2024;3(7):100950. doi:10.1016/j.jacadv.2024.100950

38. Butzner M, Cuffee Y. Telehealth Interventions and Outcomes Across Rural Communities in the United States: Narrative Review. J Med Internet Res. 2021;23(8):e29575. doi:10.2196/29575

39. Tolu-Akinnawo O, Ezekwueme F, Awoyemi T. Telemedicine in Cardiology: Enhancing Access to Care and Improving Patient Outcomes. Cureus. Published online June 21, 2024. doi:10.7759/cureus.62852

40. Azizi Z, Broadwin C, Islam S, et al. Digital Health Interventions for Heart Failure Management in Underserved Rural Areas of the United States: A Systematic Review of Randomized Trials. J Am Heart Assoc. 2024;13(2):e030956. doi:10.1161/JAHA.123.030956

41. Klee D, Pyne D, Kroll J, James W, Hirko KA. Rural patient and provider perceptions of telehealth implemented during the COVID-19 pandemic. BMC Health Serv Res. 2023;23(1):981. doi:10.1186/s12913-023-09994-4

42. Gruca TS, Pyo T, Nelson GC. Providing Cardiology Care in Rural Areas Through Visiting Consultant Clinics. J Am Heart Assoc. 2016;5(7):e002909. doi:10.1161/JAHA.115.002909

43. Vaughan L, Edwards N. The problems of smaller, rural and remote hospitals: Separating facts from fiction. Future Healthc J. 2020;7(1):38–45. doi:10.7861/fhj.2019-0066

44. Billi JE, Pai CW, Spahlinger DA. The effect of distance to primary care physician on health care utilization and disease burden: Health Care Manage Rev. 2007;32(1):22–29. doi:10.1097/00004010-200701000-00004

